# Should emotional labour be a critical component in healthcare workforce modelling?

**DOI:** 10.1101/2025.10.16.25338159

**Authors:** Cidalia Eusebio, Alison Leary, Joseph Brennan, Sue Pavord, Josh Wright, Geoffrey Punshon, Adéle Stewart-Lord

## Abstract

Healthcare work is emotionally intensive, yet emotional labour is overlooked in workforce modelling, with significant impact in terms of burnout and staff retention. The aim of this study was to examine emotional labour expenditure in a multi-professional group working in a large medical specialism across the UK to establish how this type of labour should be considered in workforce/labour modelling.

The Discrete Emotional Labour Scale was administered to multidisciplinary members of a professional society working in haematology across the UK. Principal Component Analysis was used to understand the ways in which different emotion groups interact and contribute to emotional labour. Mann-Whitney U tests were conducted to explore differences along these principal components between cohorts separated by profession, pay grade and years of service.

The findings reveal emotional labour demand can be assessed and integrated into workforce modelling. Suppressing negative and faking positive emotions significantly impact labour expenditure. Emotional labour demands vary across professional groups, pay, and years of service. Lower pay bands in clinical roles show different emotional patterns compared to higher pay bands with managerial responsibilities. Early-career professionals displayed distinct emotional labour responses compared to experienced colleagues. This highlights the need to include emotional labour demand in workforce modelling.

## INTRODUCTION

Emotional labour - the process of managing and often suppressing one’s emotions to meet professional expectations - is a significant yet unexplored aspect of healthcare workforce modelling (Riley, 2016). It has been linked to both psychological health and employee productivity, with previous studies identifying its role in burnout, stress, and emotional exhaustion among healthcare professionals (Indregard, 2018; Hu, 2023). Despite its recognised impact, emotional labour remains poorly conceptualised and operationalised in workforce planning.

The Discrete Emotional Labour Scale (DEELS) was developed by Glomb and Tews, and it is a validated tool for estimating emotional labour (Glomb, 2004). It offers an opportunity to bridge this gap by quantifying its components and integrating them into workforce models. Incorporating emotional labour expenditure into workforce planning could enhance job satisfaction, reduce burnout, and better reflect the true labour expenditure of healthcare professionals. This is particularly relevant, where high attrition rates and increased workloads pose significant challenges to workforce sustainability.

In a recent English National Health Service (NHS) survey, worsening staffing levels, increased workloads, and significant physical and mental exhaustion were reported, with many staff underreporting their worries due to fear of repercussions (NHS England, 2024). Current NHS workforce planning relies heavily on demand and capacity modelling rooted in Taylorism, which uses activity-based metrics such as appointment counts or surgeries as proxies for labour demand (NHS Staff Survey, 2024; NHS, 2019). This approach overlooks critical aspects of labour, such as emotional, cognitive and relational dimensions, leading to an underestimation of the actual workforce capacity needed to do the work required (Jackson, 2021).

A literature search was conducted using databases such as PubMed, Scopus, and Google Scholar, alongside references from relevant journals, focusing on emotional labour. The search included studies published up to June 2023 in English. Key search terms included “emotional labour,” “healthcare workforce,” “burnout,” “stress,” and “workforce modelling.” We selected studies who theoretically described or measured emotional labour in healthcare and non-healthcare contexts and its impact on the workforce. The Discrete Emotional Labour Scale (DEELS) was identified as a validated instrument for quantifying emotional labour (Glomb, 2004). Emotional labour, defined as the process of managing and often suppressing one’s emotions to meet professional expectations, is a recognised but underexplored aspect of healthcare workforce planning (Glomb, 2004). Previous studies have linked emotional labour to burnout, stress, and emotional exhaustion among healthcare professionals (Indregard, 2018; Hu, 2023). However, despite its impact, emotional labour has not been adequately conceptualised or operationalised in workforce models. Existing workforce planning in the NHS, for example, primarily relies on activity-based metrics and broad supply and demand metrics (NHS England, 2024). This gap often results in the underestimation of workforce capacity and its emotional demands. Moreover, challenges like high attrition rates and increased workloads among NHS staff and across the globe exacerbate the need for a more comprehensive approach to workforce planning accounting for emotional labour.

This study represents an initial step towards building effective emotional labour tools which can be integrated in workforce modelling. Future research should explore emotional labour across diverse healthcare settings and professional groups, as well as longitudinally assess the impact of interventions on workforce outcomes. Such efforts will ensure emotional labour is systematically addressed to enhance workforce sustainability and well-being, particularly in high-demand and complex specialties like haematology.

This study addresses these gaps by exploring how emotional labour can be integrated into workforce modelling in healthcare. It examines the relative impact of individual emotions on healthcare professionals through PCA. Additionally, the study investigates how these components vary across different professions, pay grades, and years of experience. Identifying differences between groups could help refine workforce modelling and the demand for different types of labour. This approach provides a foundation for more holistic and accurate workforce planning capturing the complexity of “work as done” rather than relying solely on traditional activity - based metrics.

## MATERIALS AND METHODS

### Design

Data was gathered using the DEELS instrument as part of a larger workforce modelling project from December 2023 to February 2024. Data was collected across England, Scotland, Wales, and Northern Ireland.

Inclusion criteria included healthcare professionals working in the field of haematology in the United Kingdom who were members of the British Society of Haematology (BSH). Exclusion criteria were those not working in haematology clinical roles with the UK.

### Participants

A convenience sample of 550 participants completed the survey. Of those, 38 were excluded as they did not meet the inclusion criteria: one did not give consent, twenty were practising abroad, and seventeen responded ‘Other’ to the profession question. ‘Other’ professionals were included in demographics data but not on the PCA and Mann-Whitney U test analysis. This resulted in a sample of 512 participants. Sample size was based on past studies that found similar numbers effective in recovering population factors. Research indicates sample needs vary with the complexity of relationships between variables and factors (Fabrigar, 2012; Watkins, 2018). Studies such as MacCallum et al suggest adequate sample sizes can account for these variables’ characteristics rather than relying solely on fixed ratios, making 512 participants sufficient for robust factor recovery in this context (MacCallum, 2001). Additionally, other studies have explored professionals working in different industry groups, supporting the generalizability of these standards across populations.

### Instruments

The purpose of the DEELS instrument is to conceptualise emotional labour, it has undergone validation studies and was validated in the original study using a sample size of 421 (Glomb, 2004). DEELS characterises emotional labour by the expression or lack thereof, whether felt or not, to be consistent with the “display rules” of an organisation (Glomb, 2004).

The DEELS instrument utilises three subscales: genuine expression, fake expression and suppression. Each of these subscales has fourteen discrete emotions, represented by six categories. Those are the Love category (Liking and Concern), Joy category (Enthusiasm, Happiness, and Contentment), Anger Category (Anger, Aggravation, and Irritation), Sadness Category (Distress and Sadness), Fear Category (Fear and Anxiety) and Hate Category (Hate and Disliking). As a result, the survey explored 42 emotions in total.

### Variables

Associations with professional categories, pay grade, and years of experience were tested with DEELS subscales and components.

### Analyses

Analysis was conducted using Wolfram Language (version 14.1) and Python (version 3.11.5) with the following libraries: NumPy, Pandas, Matplotlib, Seaborn, Scikit-learn, and FactorAnalyzer.

PCA was conducted to extract components that explained the most variance in the survey data across all 42 emotion categories.

The aim of the study was not to develop a new tool but rather to analyse the emotions that contributed most to these PC. By examining the explained variance and specific contributions for each component, it is possible to identify how different emotions—whether genuine, fake, or suppressed, positive or negative—interact to shape the experience of emotional labour in healthcare. These findings offer an insight into the complex dynamics of emotional regulation and expression within the workplace, enabling deeper understanding of the differences between cohorts which can then be used to impact on workforce modelling.

Finally, the Mann-Whitney U test was used to detect significant differences – at the 5% level – between professionals, pay grades and years of service along the PC of emotional labour.

## RESULTS

### Demographics

Of the 529 participants of the study, 67% were female and 32% male. Age distribution was primarily between 35 to 54 years, accounting for 65% of respondents. Geographically, participants were spread across various regions, with the highest representation from London (14%) and the South-West (13%). Ethnically, 69% identified as White British, with the remaining participants representing diverse backgrounds. Professionally, physicians comprised 61% of the cohort, followed by registered nurses at 18%. In terms of pay grade, 45% were consultant physicians, and 18% were in Band 7 positions.

Experience levels varied, with 63% having over 10 years in their respective fields. For further details see Supplementary Material Table 1.

### DEELS Groupings

In their original paper, Glomb & Tews identified six groups as “positive” and “negative” constituents of the 14 emotions in each of the three sub-scales: genuine, fake, and suppressed. These six groups were deemed appropriate based on a factor analysis of their data (Glomb, 2004).

Table 1 presents summary statistics of these characteristics of Emotional Labour expenditure in the healthcare workforce. Participants genuinely express emotions deemed “positive”, on average a few times a week (Median = 3.20; IQ = 2.60-3.60). Genuine and suppressed “negative” emotions are typically expressed a few times a month. For the fake “positive” and fake “negative” groups, participants report a lower frequency of expression on average.

**Table 1:** Median of Healthcare Professionals DEELS.

| Variable | Subgroup | Median (IQ) |
| --- | --- | --- |
| **DEELS**<br>**Emotional Labour** | Genuine Positive | 3.20 (2.60-3.60) |
|  | Genuine Negative | 1.89 (1.56-2.33) |
|  | Suppressed Positive | 1.4 (1-2) |
|  | Suppressed Negative | 1.89 (1.44-2.56) |
|  | Fake Positive | 1.60 (1-2.4) |
|  | Fake Negative | 1.0 (1-1) |

### Principal Component Analysis

This involved fitting an ellipsoid to the responses of the 42 emotion category questions, and using Singular Value Decomposition (SVD) to find the eigenvectors which represent the directions of the ellipsoid axes. Each eigenvector has 42 elements, representing the contribution of each emotion from the original co-ordinate system, i.e. in terms of the 42 emotions from DEELS.

Principal components are the projections of the data on these eigenvectors/ellipsoid axes. The j-th PC is therefore given by the dot product of the centered data and the j-th ellipsoid axis, where is a column vector (See Supplementary Material Figure 1).

Finding the emotions making the greatest contribution to the “most major” ellipsoid axes directions gives us novel groupings that are not immediately obvious.

The six PC that explain the most variance – have the greatest eigenvalues – are shown in Table 2, as well as the specific emotions having the highest contributions to the corresponding ellipsoid axis. The explained variance drops off rapidly after the first PC, which explains 31% of the variance. Three PC are required to explain ∼50% of the variance, and six are required to explain ∼66%.

**Table 2:** The six components which explain the most variance across all DEELS categories, and their greatest contributors.

|  | Variance Explained (%) | Key Loadings (Positive) >0.25 | Key Loadings (Negative) <-0.25 | Interpretation |
| --- | --- | --- | --- | --- |
| **PC 1** | 30.92% | Suppressed Aggravation,<br>Suppressed Fear,<br>Suppressed Distress,<br>Suppressed Sadness,<br>Suppressed Anxiety,<br>Suppressed Hate,<br>Suppressed Dislike,<br>Suppressed Anger,<br>Suppressed Concern | N/A | Most Significant Component:<br>Reflects the tendency to suppress distressing or uncomfortable emotions. This is indicative of emotional labour strategies employed to maintain professionalism or avoid conflict. |
| **PC 2** | 9.74% | Fake Happiness,<br>Fake Enthusiasm,<br>Fake Liking,<br>Fake Contentment | Suppressed Enthusiasm,<br>Suppressed Liking,<br>Suppressed Happiness,<br>Suppressed Contentment | It suggests an emotional labour strategy where individuals' fake positivity while simultaneously suppressing authentic positive feelings, likely to meet social or professional expectations. |
| **PC 3** | 8.55% | Genuine Happiness,<br>Genuine Liking,<br>Genuine Contentment,<br>Genuine Enthusiasm | N/A | Reflects authentic expression of positive emotions, suggesting alignment between internal feelings and external expressions. |
| **PC 4** | 7.73% | Suppressed Happiness,<br>Suppressed Contentment,<br>Suppressed Enthusiasm,<br>Suppressed Liking | N/A | Suggests a tendency to suppress positive emotions, potentially in contexts where positivity may seem inappropriate or unwelcome. |
| **PC 5** | 5% | Fake Enthusiasm,<br>Fake Happiness,<br>Fake Liking | Genuine Aggravation,<br>Genuine Contentment,<br>Genuine Anger,<br>Genuine Dislike,<br>Genuine Irritation | Suggests a dichotomy where fake positive emotions mask genuine negative emotions. This highlights emotional dissonance, which may lead to inauthentic interactions or emotional exhaustion. |
| **PC 6** | 4.31% | Suppressed Hate,<br>Suppressed Fear | Suppressed Irritation | This component captures the suppression of certain intense negative emotions (Hate, Fear) while indicating less suppression of milder negative feelings like irritation. It may suggest varying levels of suppression based on the intensity or appropriateness of the emotion in a given context. |

Emotional labour is a complex construct, it involves different emotions, each contributing uniquely to the overall experience. These emotions coexist and interact, with their impact varying based on how individuals respond to their environment.

Principal Component 1 (31% Variance Explained) is the most significant, capturing the impact suppressing negative emotions in this cohort has on their emotional labour, drawing attention to the strategies health professionals use to maintain professionalism or avoid conflict. Component 2 (9.74%) captures the interplay between fake positive expressions and suppressed genuine positivity, reflecting efforts to project outward positivity while concealing authentic feelings. Component 3 (8.55%) emphasises the alignment of genuine positive emotions with external expressions, while Component 4 (7.73%) reveals a tendency to suppress positivity in contexts where it might seem inappropriate. Together, these components illustrate how individuals manage both positive and negative emotions under varying demands.

Component 5 (5% Variance Explained) highlights emotional dissonance, where fake positivity masks genuine negative emotions like anger and irritation, potentially leading to inauthentic interactions and emotional exhaustion. Finally, Component 6 (4.31%) reflects the suppression of intense emotions like hate and fear, with less control over milder feelings such as irritation.

### Differences in weights of Principal Component Scores in Professions, Pay Scale and Years of Service Cohorts

This study shows differences in how various professions, pay grades, and years of experience influence emotional processing and emotional labour components. These findings help to understand the way in which in different roles and levels of seniority impact emotional dynamics in the workplace.

Significant differences in emotional labour were observed across professional groups. Professionals, such as nurses, nurse practitioners, allied health professionals, clinical scientists, and physician associates, exhibited distinct patterns of emotional processing compared to other staff such as Nursing Associates. Variations were also noted between physicians and registered nurses (PC2, PC3 & PC4), as well as between physician associates and other staff, highlighting unique emotional labour experiences among these groups. These findings highlight the influence of professional roles on how different emotions are processed in healthcare settings.

Emotional labour varies across pay grades, with distinct differences observed between lower and higher pay bands in NHS roles. Professionals in higher pay grades, such as Band 8a and consultants, experience emotional labour differently, likely due to increased responsibilities or leadership roles, compared to those in lower pay bands like Bands 5, 6, and 7. Mid-level grades (Bands 6 and 7) also differ from senior grades (Band 8a and above), perhaps reflecting shifts from clinical to managerial duties and the differences in emotional regulation needed.

Years of experience significantly influence emotional labour (PC 2, 4, 5), with differences between early-career (1-3 years), mid-career (7-10 years), and highly experienced professionals (over 10 years) (Table 3). Early-career staff manage emotions differently from their more experienced colleagues, suggesting approaches to specific emotional labour components evolve with career progression, reflecting greater experience and coping skills.

**Table 3:** Emotional Labour Components and their weight in years of experience.

| **PC** | **YEARS OF EXPERIENCE PAIRING** |  | **P-VALUE** |
| --- | --- | --- | --- |
| **PC1** |  |  |  |
| **PC2** | 4–6 years | over 10 years | 0.026 |
| **PC3** |  |  |  |
| **PC4** | Less than a year | 4–6 years | 0.031 |
|  | 1–3 years | 4–6 years | 0.022 |
|  | 4–6 years | Would rather not say | 0.020 |
|  | over 10 years | Would rather not say | 0.024 |
| **PC5** | Less than a year | Would rather not say | 0.030 |
|  | 1–3 years | 4–6 years | 0.022 |
|  | 1–3 years | over 10 years | 0.0089 |
| **PC6** |  |  |  |

### Differences in Emotions Group Scores of Professions, Pay Scale and Years of Service Cohorts

Analysis of emotional expressions revealed differences across professional groups, pay scales, and experience levels. Registered nurses and physicians differed in expressing genuine positive emotions, with variations also noted between staff in lower and middle pay bands (Bands 5-7) and senior roles (Band 8A and above) (Table 4).

**Table 4:** Differences in Emotions by profession.

| ***Emotion Group*** | ***Profession Pairing*** |  | ***p-value*** |
| --- | --- | --- | --- |
| *Genuine Positive* | Physician | Registered Nurse | 0.0058 |
| *Genuine Negative* | Registered Nurse | Clinical Scientist | 0.049 |
|  | Nursing Associate | Clinical Scientist | 0.0089 |
|  | Physician Associate | I am unregistered | 0.028 |
| *Fake Positive* | Physician | Physician Associate | 0.017 |
|  | Registered Nurse | I am unregistered | 0.043 |
|  | Pharmacist | I am unregistered | 0.012 |
|  | AHP registered with HCPC | I am unregistered | 0.018 |
|  | Pre-registration (trainee/apprentice) | Physician Associate | 0.043 |
|  | Clinical Scientist | Physician Associate | 0.030 |
|  | Physician Associate | I am unregistered | 0. |
| *Fake Negative* |  |  |  |
| *Suppressed Positive* | AHP registered with HCPC | Clinical Scientist | 0.031 |
| *Suppressed Negative* | Clinical Scientist | Physician Associate | 0.024 |
|  | Physician Associate | I am unregistered | 0.036 |

Genuine negative emotions varied between clinical scientists and both registered nurses and nursing associates, perhaps reflecting differences in patient-facing versus laboratory roles.

Early-career professionals (1-3 years) differed from mid-career staff (7-10 years) in expressing fake negative emotions, suggesting a learning curve in workplace expectations. Suppressed positive emotions were more common among senior staff compared to mid-level roles. Fake positive emotions showed the broadest associations, with higher pay bands (7A, 8A-D) and retired/non-NHS staff likely influenced by pressures for professional decorum in senior or permanent positions (Table 5).

**Table 5:**
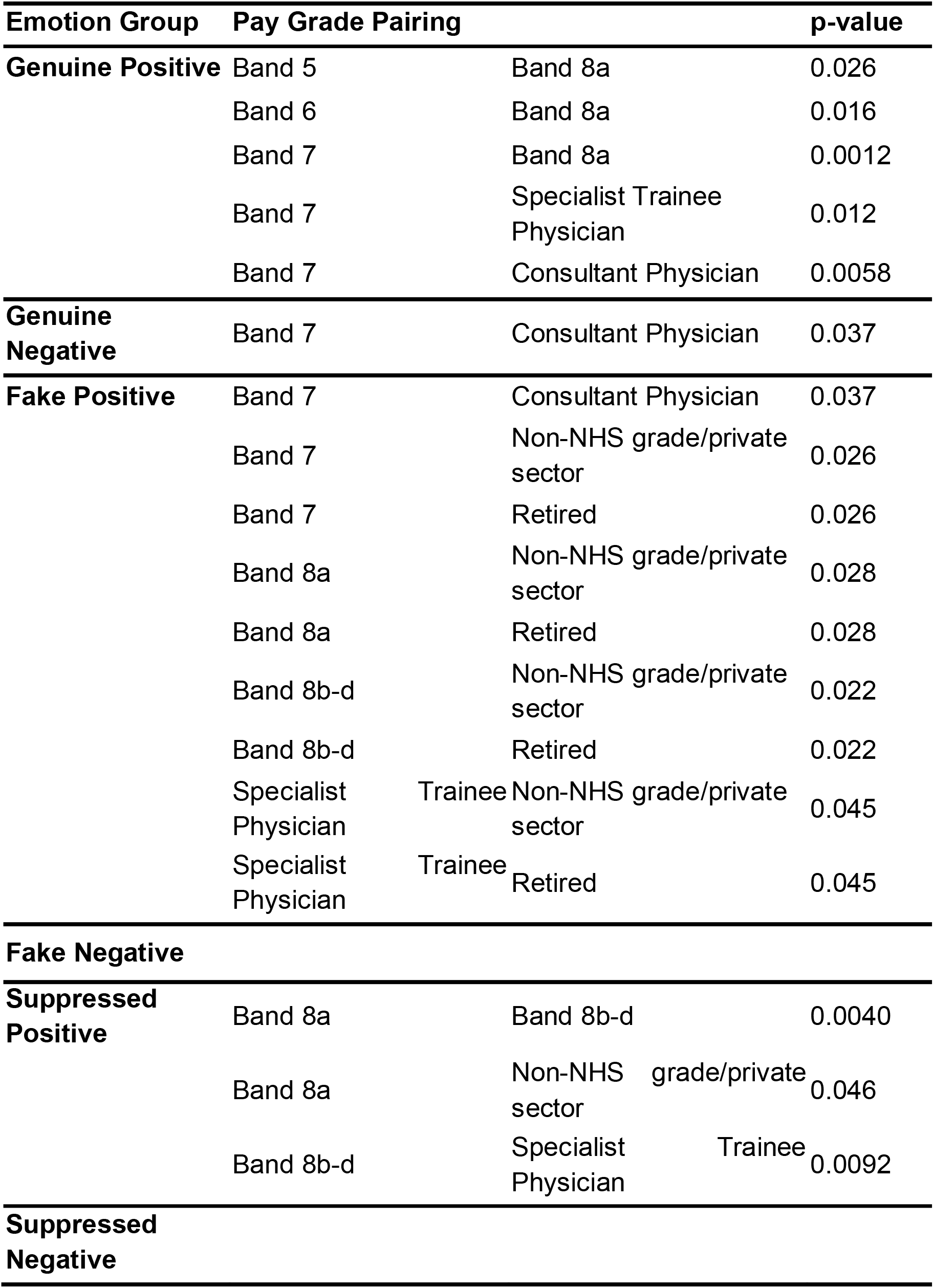
Differences in Emotions by Pay Grade.

## DISCUSSION

Emotional labour is not a uniform concept. It involves a complex interplay of emotions, each influenced by distinct weights and interactions. These emotions coexist and interact, shaping the emotional demands faced by individuals and their roles. While it is possible to create a composite score to represent emotional labour and integrate it into workforce models for the overall healthcare workforce, this score must account for variations in how different emotions—genuine, suppressed, fake, positive, or negative—interact and influence healthcare professionals.

Importantly, this study identified statistically significant differences on how emotional labour is experienced across roles, levels of experience, and pay grades. This highlights that while a general composite score can be useful for an overall comparison, different roles, levels of experience and pay grades require distinct weights for emotional labour.

The most significant components in emotional labour for healthcare professionals are suppressing negative emotions followed by displaying fake positive emotions while simultaneously suppressing authentic positive feelings. This is concerning, as previous studies have linked these behaviours to increased risks of burnout and emotional exhaustion (Singh, 2024; Spiller, 2021).

This conflict may also be due to the professional expectations in medicine and nursing, where the ideal emotionally detached rational clinician dominates medical and nursing culture and the pressure to appear empathetic without expressing their struggles often leads to disconnection of emotions, burnout and even depression (Eriksson, 2021; Leary, 2024; Liu, 2022). This creates inner conflict for professionals, highlighting the need for better support systems to help them express their emotions safely.

While these components explain the overall variability in emotional labour, it is important to note for some individuals, other components might carry more weight. These differences could lead to varying levels of job satisfaction or emotional exhaustion and burnout depending on the person’s role and context.

When examining the impact of emotional labour across different professions, pay scales, and levels of experience, statistical differences were identified between groups in specific components of emotional labour. These findings suggest potential differences between roles in management positions versus clinical roles, as well as the influence of years of experience on the management of emotional labour.

Professionals seem to have different levels of emotional labour. Differences were observed between registered and unregistered staff (in suppressed their negative emotions), as well as between physicians and nurses. These variations appear to stem from differing levels of patient contact, clinical autonomy, and organisational expectations. A recent psychosocial work environment study, although in a different healthcare system, supports these findings, showing physicians reported higher quantitative demands, while nurses faced greater emotional demands (Gynning, 2024). Nursing assistants, on the other hand, experienced the highest imbalance between efforts and rewards (Gynning, 2024).

In contrast, previous studies have also shown nurses often face greater demands with fewer resources (Eriksson, 2021). These findings suggest emotional labour is not only shaped by clinical demands but also by the distinct extrinsic working conditions experienced by different professionals in the same context (e.g. nurses versus doctors). This highlights the need to consider role-specific challenges when estimating addressing emotional labour demands in healthcare (Leary, 2024).

Experience levels also appear to influence emotional labour. Early-career professionals face distinct challenges in managing emotional labour, specifically fake negative and suppressed negative, potentially due to limited coping mechanisms, whereas experienced professionals develop strategies over time to manage their outward facing emotions more effectively. Two studies found experienced nurses and doctors expressed more genuine emotions that align with their authentic feelings during interactions with patients differently than those in early careers (Liu, 2022; Psilopanagioti, 2012).

Higher pay grades, often tied to leadership roles, demonstrate distinct emotional labour dynamics compared to lower pay grades focused on direct patient care. Managerial roles emphasise composure and neutrality, while clinical roles require empathy and emotional adaptability during patient interactions. As professionals transition from clinical roles (pay band 5–7) to administrative or leadership roles (pay band 8A and above), they face evolving emotional regulation demands. For instance, a study on nurses found those in managerial roles score higher in surface acting, indicating a greater tendency to mask true emotions or maintain a professional façade, even when their internal feelings differ from their outward expressions. These variations highlight the need for tailored emotional regulation strategies across hierarchical levels (Liu, 2022).

In summary, emotional labour is a complex and multi-layered. Its expenditure affects healthcare professionals differently depending on their roles, the emotions they manage, and the expectations placed on them. Understanding these dynamics is crucial for creating better systems to support staff and reduce the risks of burnout and emotional strain. Integrating emotional labour into workforce models offers a promising approach to addressing the emotional complexities of healthcare work, its wellbeing and sustainability.

This study is one of the first to examine emotional labour in healthcare as a facet of workforce modelling, highlighting variations across roles, experience, and pay grades. However, several limitations must be noted. The study did not explore the directionality of associations between emotional labour, profession, pay grade, and experience. This study necessarily relied on self-reported data, which may introduce bias. Additionally, the findings are specific to haematology professionals, with a sample skewed towards senior, experienced professionals and physicians and may not generalise to other healthcare contexts. Future research should investigate emotional labour across diverse clinical specialties and explore interventions to reduce its burden. Due to the orthogonality of principal components, individual scores cannot be meaningfully combined. Future research should assess whether the six selected components adequately capture emotional labour for all respondents. Longitudinal research is needed to establish causation and assess its impact on workforce sustainability and explore the underlying reasons for variations in emotional labour across clinical, managerial and experience.

## CONCLUSION

In this study, the Discrete Emotional Labor Scale (DEELS) was used to examine the relationship between the different components of emotional labour and professions, pay grade and years of experience and how those can be used to model the healthcare workforce. Findings indicate suppressing negative emotions and faking positive ones significantly contributes to the emotional labour expenditure of healthcare professionals. However, this burden varies across professional roles, pay grades, and years of experience. These dynamics should be carefully considered when modelling the healthcare workforce to ensure a balance not only in workforce supply but also in their sustainability and workforce wellbeing. Existing evidence establishes the relationship between emotional labour, and its impact on staff retention, burnout, and emotional exhaustion. These findings confirm this within healthcare, demonstrating the feasibility of quantifying emotional labour and its varying effects across roles, pay grades, and career stages. This evidence supports the development of sustainable healthcare systems by advocating for the integration of emotional labour considerations into workforce modelling. Modelling workforce considering the impact of emotional labour is critical for reducing burnout and improving retention, particularly among professionals in emotionally demanding roles or critical career stages.

## Figure Legends

**Supplementary Material Figure 1:**
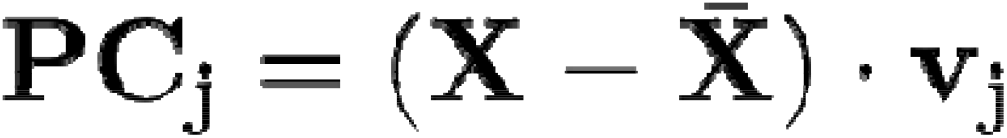
Principal component analysis formula.

## Table Legends

Supplementary Material Table 1: Demographics of participants.

## Acknowledgments

The authors wish to thank Deborah McLean-Thorne and Theresa Crossley from the British Society for Haematology and Jon Mcloon and Srihitha Ramasahm from Wolfram for their assistance with this project.

## Compliance with Ethical Standards

Ethical approval was gained through London South Bank University Ethics Panel ETH2324-0008 and ETH2324-0134

## Data Availability

Data is available from the authors upon reasonable request.

