## Supplementary material for "Should emotional labour be a critical component in healthcare workforce modelling?": Suplemental Table 1

**Supplementary Material Table 1: Demographics of participants.**

| **Variable** | **Subgroup** | **Number of participants n=529, (proportion)** |
| --- | --- | --- |
| **Gender** | Male | 169 (32%) |
|  | Female | 357 (67%) |
|  | Prefer not to say | 3 (1%) |
| **Age (years old)** | 25 to 34 | 80 (15%) |
|  | 35 to 44 | 183 (35%) |
|  | 45 to 54 | 160 (30%) |
|  | 55 to 64 | 90 (17%) |
|  | 65 to 74 | 7 (1.4%) |
|  | 75 or older | 3 (0.6%) |
|  | Prefer not to say | 6 (0.4%) |
| **Regions** | England - East | 30 (6%) |
|  | England - East Midlands | 47 (9%) |
|  | England - London | 74 (14%) |
|  | England - North East | 34 (7%) |
|  | England - North West | 54 (10%) |
|  | England - Northern and Yorkshire | 54 (10%) |
|  | England - South Central | 26 (5%) |
|  | England - South East | 34 (6%) |
|  | England - South West | 71 (13%) |
|  | England - West Midlands | 42 (8%) |
|  | Northern Ireland | 14 (3%) |
|  | Scotland | 29 (5%) |
|  | Wales | 20 (4%) |
| **Ethnicity** | White British English, Welsh, Scottish, Northern Irish or British Irish | 367 (69%) |
|  | Any other white background | 32 (6%) |
|  | Indian | 21 (4%) |
|  | Prefer not say | 14 (3%) |
|  | Asian or British Asian | 17 (3%) |
|  | African | 12 (2%) |
|  | Chinese | 8 (2%) |
|  | Irish | 12 (2%) |
|  | Others | 46 (9%) |
| **Profession** | Physician | 321 (61%) |
|  | Registered Nurse | 93 (18%) |
|  | Professional AHP registered with HCPC | 36 (7%) |
|  | Clinical Scientist | 34 (6%) |
|  | Pre-registration (e.g. apprentice) | 7 (1%) |
|  | Pharmacist | 7 (1%) |
|  | Physician Associate | 6 (1%) |
|  | I am unregistered | 4 (1%) |
|  | Nursing Associate | 3 (0.6%) |
|  | Midwife | 1 (0.4%) |
|  | Other | 17 (3%) |
| **Pay Grade** | Consultant Physician | 238 (45%) |
|  | Band 7 | 94 (18%) |
|  | Specialist Trainee Physician | 77 (15%) |
|  | Band 8a | 43 (8%) |
|  | Band 8b-d | 28 (5%) |
|  | Band 6 | 26 (5%) |
|  | Band 5 | 6 (1%) |
|  | Band 4 | 6 (1%) |
|  | Non-NHS grade/ private sector | 4 (1%) |
|  | Retired/ Would rather not say | 7 (1%) |
| **Experience in years** | Over 10 years | 334 (63%) |
|  | 7-10 years | 71 (13%) |
|  | 4-6 years | 64 (12%) |
|  | 1-3 years | 42 (8%) |
|  | Less than year | 16 (3%) |
|  | Would rather not say | 2 (1%) |
